# IDENTIFICATION OF MODIFIER GENE VARIANTS OVERREPRESENTED IN FAMILIAL HYPOMAGNESEMIA WITH HYPERCALCIURIA AND NEPHROCALCINOSIS PATIENTS WITH A MORE AGGRESSIVE RENAL PHENOTYPE

**DOI:** 10.1101/2023.11.08.23298254

**Authors:** M Vall-Palomar, J Morata, M Duran, J Torchia, R Tonda, M Ferrer, A Sánchez, G Cantero-Recasens, G Ariceta, A Meseguer, C Martinez

**Affiliations:** Renal Physiopathology Group, Vall d’Hebron Research Institute (VHIR), Vall d’Hebron Barcelona Hospital Campus, Barcelona, Spain; CNAG-CRG, Centre for Genomic Regulation (CRG), Barcelona Institute of Science and Technology (BIST), Barcelona, Spain; Statistics and Bioinformatics Unit (UEB), Vall d’Hebron Research Institute (VHIR), Vall d’Hebron Barcelona Hospital Campus, Barcelona, Spain; Universitat de Barcelona, Barcelona, Spain; Pediatric Nephrology Department, Vall d’Hebron University Hospital, Universitat Autònoma de Barcelona, Barcelona, Spain; Departament de Bioquímica i Biologia Molecular, Unitat de Bioquímica de Medicina, Universitat Autònoma de Barcelona, Barcelona, Spain; Vascular and Renal Translational Research Group. Lleida Institute for Biomedical Research Dr. Pifarré Foundation (IRBLleida), Lleida, Spain

**Author notes:** Correspondence: AM.

**Keywords:** Rare renal disease, renal tubulopathy, familial hypomagnesemia with hypercalciuria and nephrocalcinosis, phenotype modifier genes, CLDN16, CLDN19, FHHNC, whole exome sequencing

## Abstract

Inter- and intra-familial phenotypic variability is a common observation in genetic diseases. In this study we have gathered a highly unique patient cohort suffering from an ultra-rare renal disease, familial hypomagnesemia with hypercalciuria and nephrocalcinosis, with a deep clinical and genetic characterization. In this cohort, we have previously reported a high phenotypic variability between patients harbouring exactly the same mutation in homozygosis (70% of patients), even between siblings. Patients were stratified at the extremes according to their estimated glomerular filtration rate annual decline and subjected to whole exome sequencing aiming to find candidate phenotype modifier genes. The analysis pipeline applied has allowed us to find, for the first time, 17 putative modifier gene variants associated with a more aggressive renal phenotype. Our results led to a panel of genetic variants in novel candidate modifier genes which will be useful to stratify patients according to their risk of developing renal failure earlier in life and, therefore, direct them to more appropriate and personalized therapeutic options.

## 1. INTRODUCTION

Familial hypomagnesemia with hypercalciuria and nephrocalcinosis (FHHNC) is an ultra-rare autosomal recessive renal tubular disease with an incidence of <1/1.000.000 individuals, characterized by severe urinary Ca^2+^ and Mg^2+^ wasting and progression to chronic kidney disease (CKD) and renal failure^1,2^. Disease loss-of-function causing mutations have been identified in the tight junction (TJ) proteins claudin 16 (CLDN16)^3^ and 19 (CLDN19)^4^. Both proteins are co-expressed in the TJ of the thick ascending limb of Henle’s loop and form a paracellular cation-selective pore that induces a NaCl gradient and a lumen-positive transepithelial voltage driving Ca^2+^ and Mg^2+^ reabsorption^5,6^. Additionally, *CLDN19* is also expressed in the retinal epithelium and a subset of FHHNC patients with *CLDN19* mutations develop severe ocular impairment^4^. No specific therapy for FHHNC exists and patients rely only on supportive treatment (high fluid intake, dietary restrictions, magnesium salts and thiazide diuretics) attempting to delay the progression to renal failure. Kidney replacement therapy remains the only curative option in end-stage renal disease patients and renal transplant is usually required during the second to third decades of life^1,2^, causing a severe impairment on quality of life of patients from a very young age.

Major research efforts in the last decade have been directed towards identifying novel mutations in FHHNC patients^7^ and, as a result, 73 and 24 different mutations in *CLDN16* and *CLDN19*, respectively, have been described and annotated in the Human Gene Mutation Database (HGMD)^8^. Most FHHNC patients have been found to carry *CLDN16* mutations, although in the South of Europe (mainly Spain and France) *CLDN19* mutations are more prevalent and a specific *CLDN19* so-called Hispanic founder mutation (c.59G>A; p.G20D) has been described^9,10^. Remarkably, carriers of the p.G20D mutation exhibit a prominent, but yet unexplained, phenotypic variability which is observed even between homozygotic siblings^2,9,11,12^. This suggests the existence of other not-so-far-identified molecular events, such as phenotype modifier genes, that might contribute to the clinical variability beyond the causal genes. Here we present the first study focusing on the identification of phenotype modifier genes in a unique cohort of 30 FHHNC Spanish patients through whole exome sequencing (WES) analysis comparing extreme phenotypes, as it has been successfully applied to other complex Mendelian diseases^13-15^. Candidate modifier genes were then selected by a thorough prioritization strategy combining publicly available information regarding described variants associated to disease-specific phenotypic traits from the GWAS catalog together with kidney-specific expression data from the Human Protein Atlas and Genotype Tissue Expression (GTEx) databases. Our results led to a panel of genetic variants in novel candidate modifier genes which will be useful to stratify patients according to their risk of developing renal failure earlier in life and, therefore, direct them to more appropriate and personalized therapeutic options.

## 2. METHODS

### 2.1. Cohort description

This study included a cohort of 30 patients with clinical and genetic confirmation of FHHNC and stratified according to our previously reported cutoff for the annual glomerular filtration rate (eGFR) decline estimation^12^, as follows: ≥ 10 mL/min/1.73m^2^/year for the fast renal progression group; < 10 mL/min/1.73m^2^/year for the moderate-renal progression group; no eGFR decline among time (stable renal function) for the slow-renal progression group. Genetic diagnosis and clinical phenotype is thoroughly described in our previous study in the same cohort^12^.

All patients, or their caregivers, provided written informed consent before participation in the study. This study was approved by the Ethics Committee of our institution, the Vall d’Hebron Hospital (PR(AMI)280/2015). Patient records were consulted only to obtain relevant patient data. The described research adhered to the Declaration of Helsinki.

### 2.2. Whole exome sequencing

#### Library preparation and sequencing

DNA from patients was obtained using the Gentra Puregene Blood kit® (#158467, Qiagen), following manufacturers’ instructions. Exome sequencing was performed at the Centro Nacional de Análisis Genómico (CNAG-CRG, Barcelona, Spain). Paired-end multiplex libraries were prepared according to manufacturer’s instructions and enriched with the SureSelect Human All Exon v5 genome design exome kit (Agilent Technologies). Libraries were loaded to Illumina flowcells for cluster generation prior to producing 100 base read pairs on a NovaSeq6000 instrument following the Illumina protocol. Image analysis, base calling and quality scoring of the run were processed using the manufacturer’s software Real Time Analysis (RTA 1.18.66.3) and followed by generation of FASTQ sequence files with the CASAVA software.

#### Data analysis and variant calling

Reads were mapped to Human reference genome v37 with decoy sequences (ftp://ftp.1000genomes.ebi.ac.uk/vol1/ftp/technical/reference/phase2_reference_assembly_sequence/hs37d5.fa.gz) with Burrows-Wheeler Alignment tool 0.7.17 (BWA-MEM)^16^. Alignment files containing only properly paired, uniquely mapping reads without duplicates were processed using Picard 2.20^17^ to add read groups and to remove duplicates. The Genome Analysis Tool Kit (GATK 4.1.9.0)^18^ was used for local realignment and base quality score recalibration. Variant calling was done using HaplotypeCaller from GATK. Functional annotations were added using SnpEff v5 with the GRCh37.75 database^19^. Variants were annotated with SnpSift v5^20^ using population frequencies, conservation scores and deleteriousness predictions from dbNSFPv4.1a^21^. Other sources of annotations, such as gnomAD, CADD and Clinvar were also used^22,23^. Statistical analysis was performed for variants of snpEff predicted impact moderate or high and with a coverage of at least 10 reads in one sample. Association tests were performed with Rvtests’ SKAT-O^24^. Fisher Tests were additionally performed with R fisher.test package (Fisher’s Exact Test for Count Data).

### 2.3. Strategy for prioritization of candidate genes

The strategy we followed to select the most interesting candidate genes for further analysis was as follows:

1. P-value threshold <0.01
2. Genes expressed in the kidney (at gene and protein evidence) using the information obtained from the Human Protein Atlas.

Then we investigated whether significant genes obtained by SKAT-O had been previously associated with particular phenotype traits related to FHHNC. To do this, variant-trait associations were extracted from the GWAS Catalog^25^ and those showing a P-value < 1 × 10^−6^ were selected and matched to our list of SKAT-O genes. Supplementary table 2 summarizes the selected phenotypic traits that were searched in the GWAS catalog, the number of associated variants found for each trait and the number of matched genes from our SKAT-O analysis.

Finally, a second round of GWAS catalog – SKAT-O matching genes was performed including genes not expressed in the kidney. Members of the claudin family of proteins were also included.

### 2.4. Functional Analysis

To gain further insights into the functional relevance of the genes identified by SKAT-O, several approaches and tools were used, as described below. The main goal was to group the prioritized candidate genes into pathways and biological processes to further explore their potential involvement in FHHNC phenotype modulation. The statistical analysis was performed using the statistical language “R” (R version 4.2.0)^26^ and libraries from the Bioconductor Project (www.bioconductor.org).

#### g:Profiler analysis

The version of g:Profiler used for this study was e107_eg54_p17_bf42210. The parameters for the enrichment analysis were as follows. Homo sapiens (human) was chosen as the organism for the analysis. GO analyses (GO molecular function (GO-MF), GO cellular component (GO-CC), and GO biological process (GO-BP)) were carried out sequentially. The statistical domain scope was used only for annotated genes and the significance threshold was the Benjamini and Hochberg FDR threshold at < 0.05.

#### Overrepresentation analysis (ORA)

ORA of the selected genes was performed over the GO-BP database using clusterProfiler R package v4.4.1^27^. The background distribution for the hypergeometric test was all the human genes that have annotation. Gene sets were obtained from the annotation package org.Hs.eg.db v.3.15.0. A list of the enriched terms sorted by P-value and filtered by an adjusted P-value <0.25 was obtained. Results were graphically represented as a dot plot of the top 15 terms enriched with an adjusted P-value <0.25. In this plot, the size of the dots relates to the number of genes/proteins in the data that belong to that pathway, the colour of the plot refers to significance level (P-value). The terms are ordered by P-value and Gene ratio, which is the ratio between the genes/proteins in the data that belong to that term and the total number of genes/proteins in the term.

#### Summarization of ORA results

To summarize the results obtained in the enrichment analysis over GO-BP, a clustering analysis was performed using the R/Bioconductor package simplifyEnrichment v1.6.1^28^. This method allows to cluster terms using different similarity measures and clustering algorithms and visualize the results as Heatmaps. Here, terms with an adjusted P-value <0.25 were grouped based on Rel similarity measure and binary cut clustering algorithm^28^. Clusters were annotated with a word cloud representing the top 10 most frequent words found within term names in each cluster, and with the most representative term within the cluster, selected as the term with maximal number of SKAT-O genes and with the minimal P-value).

### 2.5. Statistical analysis

The biostatistics analysis was focused on the identification of gene variants associated to the FAST renal progression phenotype. Prioritized modifier candidate genes were analysed by the exact Fisher test considering patients homozygous for the reference allele (R/R), heterozygous (R/A) and homozygous for the alternative allele (A/A) among the two phenotypes (FAST *vs*. noFAST) to identify those variants that distribute differently between both phenotypes. For significant variants, the odds ratios (OR) and 95% confidence intervals were calculated applying the Haldane and Anscombe correction^29^, for the homozygous A/A or the heterozygous A/R genotypes compared to the homozygous R/R genotype.

To gain further insights on the association of prioritized variants with fast renal progression, we used eQTL data to check variant expression by genotype in 73 human kidney cortex tissues from the GTEx eQTL Dashboard. The data used for the analyses described in this manuscript were obtained from the GTEx Portal on 01/23. In addition, to find whether the distribution of all genotypes (A/A, A/R, or R/R) was the same for each group of subjects analysed (FAST, noFAST, GTEx) data was analysed by the exact Fisher’s test.

The statistical analysis was performed using the statistical language “R” (R version 4.2.0 (2022-04-22), Copyright (C) 2022 The R Foundation for Statistical Computing).

## 3. RESULTS

### 3.1. Clinical characterization of the FHHNC cohort

As previously described^12^, the cohort was composed of 30 patients from 22 unrelated families. Pathogenic *CLDN16* mutations were confirmed in 3 patients, while the remaining 27 patients carried *CLDN19* mutations (74% were homozygous for the p.G20D mutation). Patients were classified based on the annual renal function decline, following the clinical criteria previously reported by our group^12^ (figure 1, supplementary table 1).

**Figure 1.**
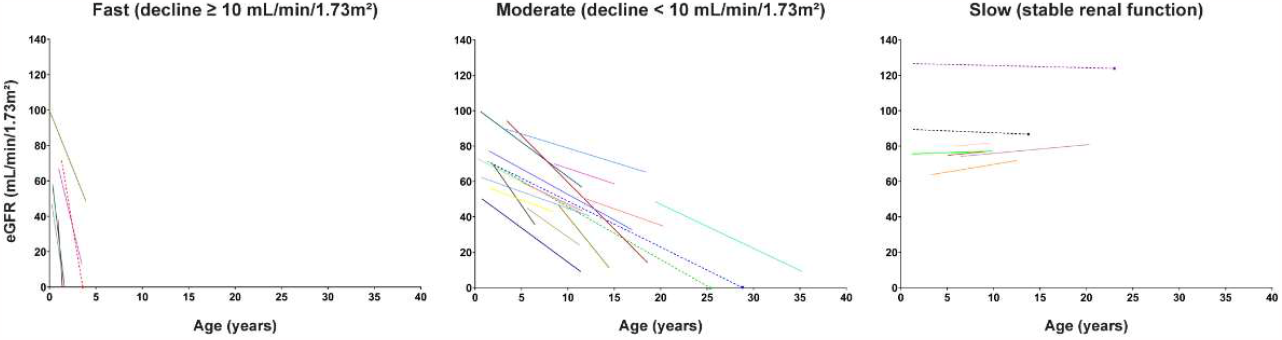
Annual renal function decline. Patterns of eGFR change over time were plotted for each patient and stratified as fast, moderate or slow according to the curve slopes determined by linear regression. Each coloured line indicates the annual eGFR loss. Dashed lines indicate patients for whom we only had one value of eGFR corresponding to the last follow-up.

Patients were then stratified in two extreme phenotypes for WES analysis, FAST vs. noFAST. The FAST group included patients (n=6) with the fastest renal function decline (figure 1), while the noFAST group (n=24) comprised patients with a moderate and stable decline in eGFR rate. Remarkably, two pairs of siblings belonging to families carrying the p.G20D in homozygosis showed different renal phenotypes (fast vs. moderate) and were classified in different groups for WES analysis. Additionally, we recruited 4 healthy controls among the non-affected siblings of patients.

### 3.2. Whole exome sequencing

To identify candidate disease-modifying genes implicated in the phenotypic heterogeneity described in our FHHNC cohort, whole exome sequencing was performed. On average, we obtained high exon coverage of 97.22x for all 30 samples demonstrating good quality of the sequence data. Of all exons, 99.87% were covered (nonzero coverage) and 98.83% were assessed by at least 10 independent reads. GATK yielded 29870 variants mapping to 11240 genes across the different samples. WES results confirmed the genetic diagnostic previously defined by Sanger sequencing for each patient. SKAT-O statistics were calculated using all called variants with moderate and high effect types (missense, splice region, splice donor, splice acceptor, stop-loss, stop-gain) which yielded 1068 significant genes potentially modifying the phenotype severity (P<0.05). Functional profiling analysis revealed that these genes were enriched in GO-BP terms related to cell-cell adhesion, response to pro-inflammatory signals (interleukin 1) and tissue regeneration; and in GO-CC terms related to plasma membrane (supplementary figure 1).

### 3.3. Candidate gene prioritization identifies phenotype modifier variants that are associated to a faster progression to end-stage renal disease in FHHNC patients

The SKAT-O genes were filtered by selecting those with a P-value <0.01 and described to be expressed in the human kidney in the Human Protein Atlas database, at both RNA and protein evidence level. These filters reduced the candidate gene list to 165 genes.

As a second step to further prioritize potential modifier genes, we searched the GWAS catalog database to identify gene variants associated to FHHNC specific phenotypic traits. Those specific traits were selected based on the renal phenotype described in our patient cohort (annual glomerular filtration rate decline) plus the wide range of common FHHNC symptoms which include polyuria-polydipsia, failure to thrive, recurrent urinary tract infections, hypercalciuria, nephrocalcinosis, hypomagnesemia, and hyperparathyroidism^2,30,31^. In addition, we also included traits related to the ocular impairment described for some carriers of *CLDN19* mutations. Supplementary table 2 summarizes the selected phenotypic traits, the number of associated variants found in the GWAS catalog and the number of matched genes from our SKAT-O analysis after filtering by P<0.01 and kidney expression. As a result, the list of potential candidates for further analysis was reduced to 44 candidate modifier genes. We performed a second round of data matching between SKAT-O genes and GWAS catalog data including genes not expressed in the kidney in order to not exclude the possibility that genes from other tissue and/or cell types may have an impact on FHHNC progression. This second data-matching analysis identified only another 5 candidates. Additionally, as the claudin family of proteins are highly involved in the physiopathology of FHHNC, we included in the list of selected candidates the gene *CLDN17* that was significantly associated (P<0.01) to the FAST phenotype. Applying the described strategy for candidate prioritization, we finally selected 175 variants mapping to 50 genes for further analysis (table 1).

**Table 1.** List of 50 prioritized candidate modifier genes.

| Gene ID | SKATO<br>P-value | Kidney<br>expression | Variants in<br>FHHNC | Variants in<br>GWAS catalog | GWAS catalog main trait |
| --- | --- | --- | --- | --- | --- |
| *OSBPL6* | 0.0002 | Yes | 3 | 1 | High myopia |
| *HLCS* | 0.0004 | Yes | 3 | 1 | Estimated glomerular filtration rate |
| *NFU1* | 0.0006 | Yes | 1 | 1 | Calcium levels |
| *NINJ1* | 0.0008 | Yes | 1 | 2 | Estimated glomerular filtration rate |
| *DMD* | 0.001 | Yes | 5 | 1 | Renal system measurement |
| *IL12RB1* | 0.002 | Yes | 5 | 2 | Estimated glomerular filtration rate |
| *ANKLE2* | 0.002 | Yes | 7 | 1 | High myopia |
| *DOCK9* | 0.002 | Yes | 4 | 6 | Heel bone mineral density / low myopia |
| *CEP89* | 0.003 | Yes | 5 | 5 | Estimated glomerular filtration rate |
| *MAST2* | 0.003 | Yes | 7 | 2 | Serum creatinine levels / Serum 25-Hydroxyvitamin D levels |
| *HPS5* | 0.003 | Yes | 2 | 1 | Renal system measurement |
| *CLDN17* | 0.003 | No | 1 | 0 | NA |
| *SIN3A* | 0.003 | Yes | 1 | 4 | Estimated glomerular filtration rate |
| *RGS19* | 0.003 | Yes | 1 | 1 | Estimated glomerular filtration rate |
| *CMAHP* | 0.003 | No | 1 | 1 | Creatinine levels in ischemic stroke |
| *TXNL1* | 0.003 | Yes | 1 | 1 | 6-month creatinine clearance change response to tenofovir in HIV |
| *SLC35F1* | 0.003 | No | 1 | 1 | Heel bone mineral density |
| *GRAMD1A* | 0.003 | Yes | 1 | 1 | Calcium levels |
| *EIF1AD* | 0.003 | Yes | 1 | 1 | Renal System Measurement |
| *GLI2* | 0.003 | Yes | 3 | 1 | Estimated glomerular filtration rate in coronary artery disease and impaired kidney function |
| *SPG7* | 0.003 | Yes | 3 | 2 | Heel bone mineral density |
| *HHAT* | 0.003 | Yes | 4 | 1 | High myopia |
| *MICALL1* | 0.004 | Yes | 3 | 2 | Heel bone mineral density |
| *PLEKHG1* | 0.005 | Yes | 4 | 2 | Retinal drusen |
| *PMFBP1* | 0.005 | Yes | 4 | 3 | Urinary metabolite measurement |
| *SLC9A3* | 0.005 | Yes | 1 | 2 | Estimated glomerular filtration rate |
| *GALP* | 0.005 | No | 1 | 1 | Microalbuminuria |
| *CCDC57* | 0.005 | Yes | 10 | 1 | Renal system measurement |
| *RHPN2* | 0.005 | Yes | 4 | 12 | Bone mineral density / macular thickness |
| *UNC13C* | 0.006 | No | 2 | 2 | Retinal drusen / proteinuria in preeclampsia |
| *RBL2* | 0.006 | Yes | 5 | 1 | Estimated glomerular filtration rate |
| *NADSYN1* | 0.006 | Yes | 3 | 14 | Vitamin D levels |
| *ITGA1* | 0.006 | Yes | 5 | 2 | Heel bone mineral density |
| *PITPNM2* | 0.008 | Yes | 2 | 2 | Calcium levels / macular thickness |
| *SORT1* | 0.008 | Yes | 3 | 2 | Estimated glomerular filtration rate |
| *DLG5* | 0.008 | Yes | 5 | 1 | Retinal detachment or retinal break / macular thickness |
| *AGBL2* | 0.008 | Yes | 2 | 1 | Calcium levels |
| *CGNL1* | 0.008 | Yes | 5 | 14 | Estimated glomerular filtration rate / macular thickness |
| *CACNA1S* | 0.008 | No | 8 | 2 | Estimated glomerular filtration rate |
| *MLXIPL* | 0.008 | Yes | 4 | 8 | Renal system measurement / Calcium levels |
| *MADD* | 0.009 | Yes | 4 | 1 | Chronic kidney disease |
| *NPHS1* | 0.009 | Yes | 5 | 3 | Estimated glomerular filtration rate / Serum 25-Hydroxyvitamin D levels |
| *ZNF607* | 0.009 | Yes | 3 | 3 | Estimated glomerular filtration rate / Renal system measurement |
| *GPRIN3* | 0.01 | Yes | 6 | 1 | Urinary calcium excretion |
| *CD86* | 0.01 | Yes | 1 | 8 | Calcium levels |
| *SPATA20* | 0.01 | Yes | 3 | 3 | Estimated glomerular filtration rate / bone mineral density |
| *SLC17A1* | 0.01 | Yes | 1 | 16 | Renal system measurement |
| *MPP7* | 0.01 | Yes | 1 | 14 | Bone mineral density / macular thickness |
| *CLDN8* | 0.01 | Yes | 2 | 1 | Renal system measurement |
| *PLIN4* | 0.01 | Yes | 17 | 2 | Estimated glomerular filtration rate |

To uncover the biological significance of the prioritized genes, an overrepresentation analysis (ORA) was performed using the GO-BP followed by clustering analysis to summarize the results obtained (summORA). Among the top 15 terms identified by ORA, those related to apical junction regulation and assembly, cell adhesion and immune-related terms were the most significant biological processes associated to our 50 phenotype modifier candidates (figure 2A). After clustering by similarity the enriched GO-BP terms obtained with an adjusted P-value < 0.25, 12 clusters were identified (figure 2B). Both the most representative term found in each cluster and the 10 most frequent words within them, highlighted apical junction regulation and assembly, immune-related terms, ionic transport, protein modifications (mainly phosphorylation), cellular responses to different stimulus, and cell cycle regulation as the most significant biological processes enriched in the FHHNC prioritized modifier genes.

**Figure 2.**
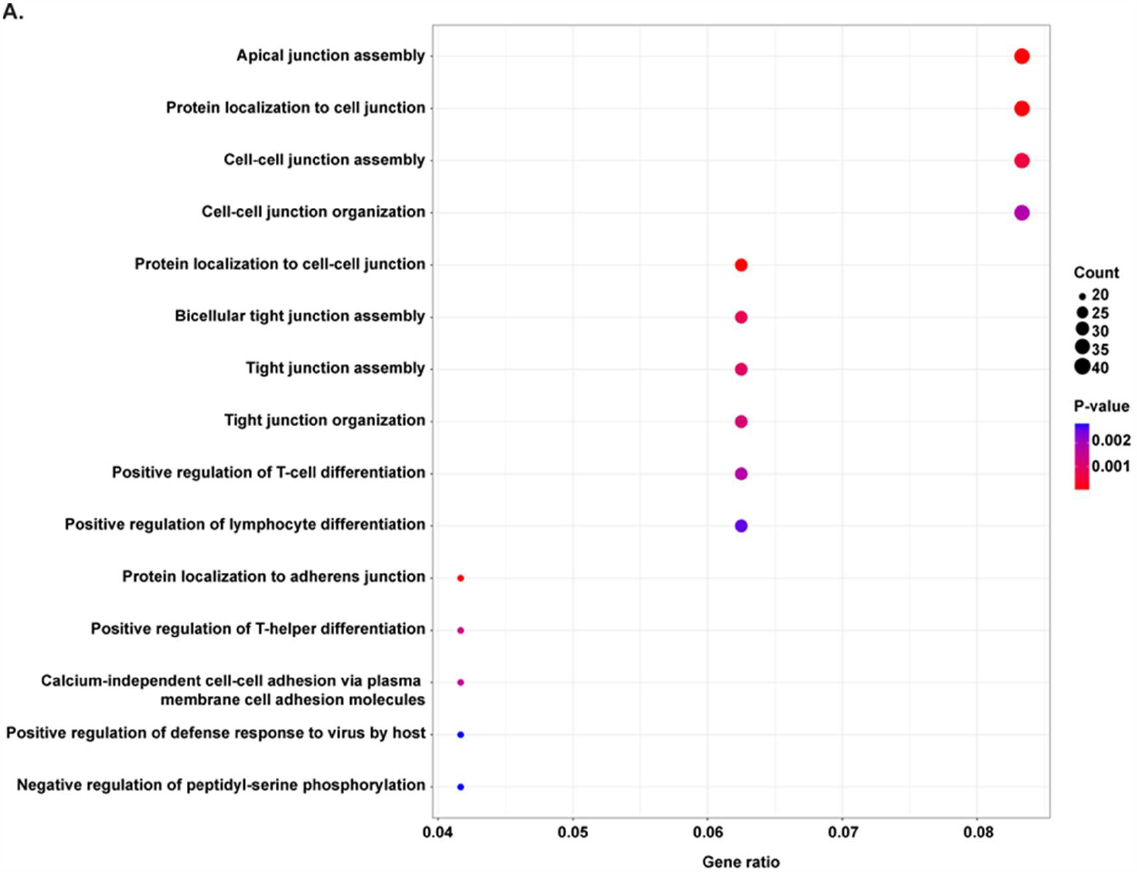
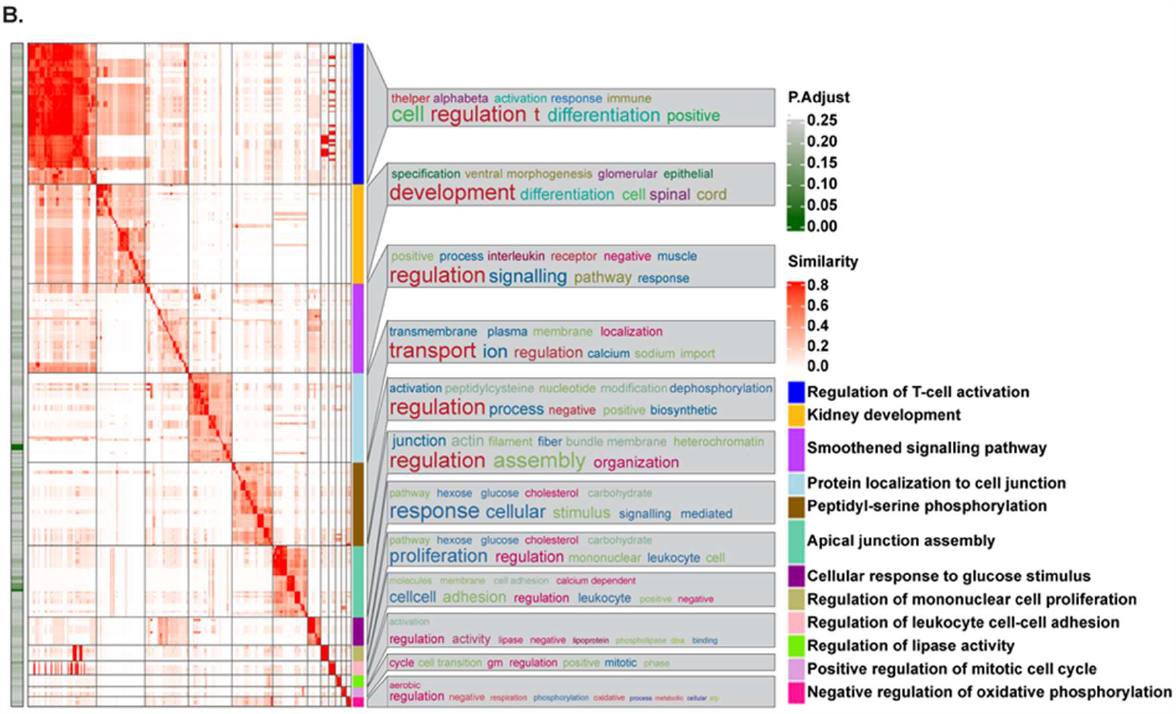
Overrepresentation analysis of the 50 prioritized candidate modifier genes in FHHNC patients. **A**. Dot plot showing the top 15 most significant biological processes overrepresented. The size of the dots relates to the number of genes/proteins in the data that belong to that pathway, the colour of the plot refers to significance level (P-value). The terms are ordered by P-value and Gene ratio, which is the ratio between the genes/proteins in the data that belong to that term and the total number of genes/proteins in the term. **B**. Heatmap of clustering analysis summarizing overrepresentation results. Clusters are annotated with the most representative term within each cluster (the term with the lowest P-value and the higher number of candidate genes) and with a word cloud representing the top 10 most frequent words found within term names in each cluster. The heatmap shows the terms found within each list, coloured according to their adjusted P-value (P.Adjust).

Since SKAT-O does not provide any information regarding whether the candidate genes confer a risk or a protective effect, we took a complementing approach by performing Fisher’s exact test and calculating odds ratios for each individual variant identified of SKAT-O prioritized candidate genes. Nine single nucleotide variants (SNVs) in homozygosis and 9 SNVs in heterozygosis were significantly associated to renal progression phenotypes (Fisher P-value <0.05; Odds ratio ≠1) considering the alternative allele both in homozygosis and in heterozygosis (figure 3, supplementary table 3). From those, the alternative allele in homozygosis for SNVs mapping to genes *CD86, CEP89, GALP, IL12RB1* and *NFU1* was associated to the FAST phenotype; while rs228406 (*DMD*) and rs8065903 (*SPATA20*) were associated to the noFAST phenotype (figure 3A). Additionally, the alternative allele in heterozygosis for SNVs mapping to genes *AGBL2, ANKLE2, CLDN17, DMD, HSP5, IL12RB1, ITGA1* and *ZNF607* was associated to the FAST phenotype; while rs8065903 (*SPATA20*) was associated to the noFAST phenotype (figure 3B). Variants in these genes are annotated in the GWAS catalog associated to Ca^2+^ levels (*CD86, NFU1, AGBL2*), eGFR (*CEP89, IL12RB1, SPATA20, ZNF607*), blood urea nitrogen levels (*DMD, HPS5, ZNF607*), microalbuminuria (*GALP*), bone mineral density (*ITGA1*) and high myopia (*ANKLE2*) (table 1). In addition, most of these genes were found in clusters related to immune system activity and regulation of apical junctions in the summORA analysis (supplementary table 4).

**Figure 3.**
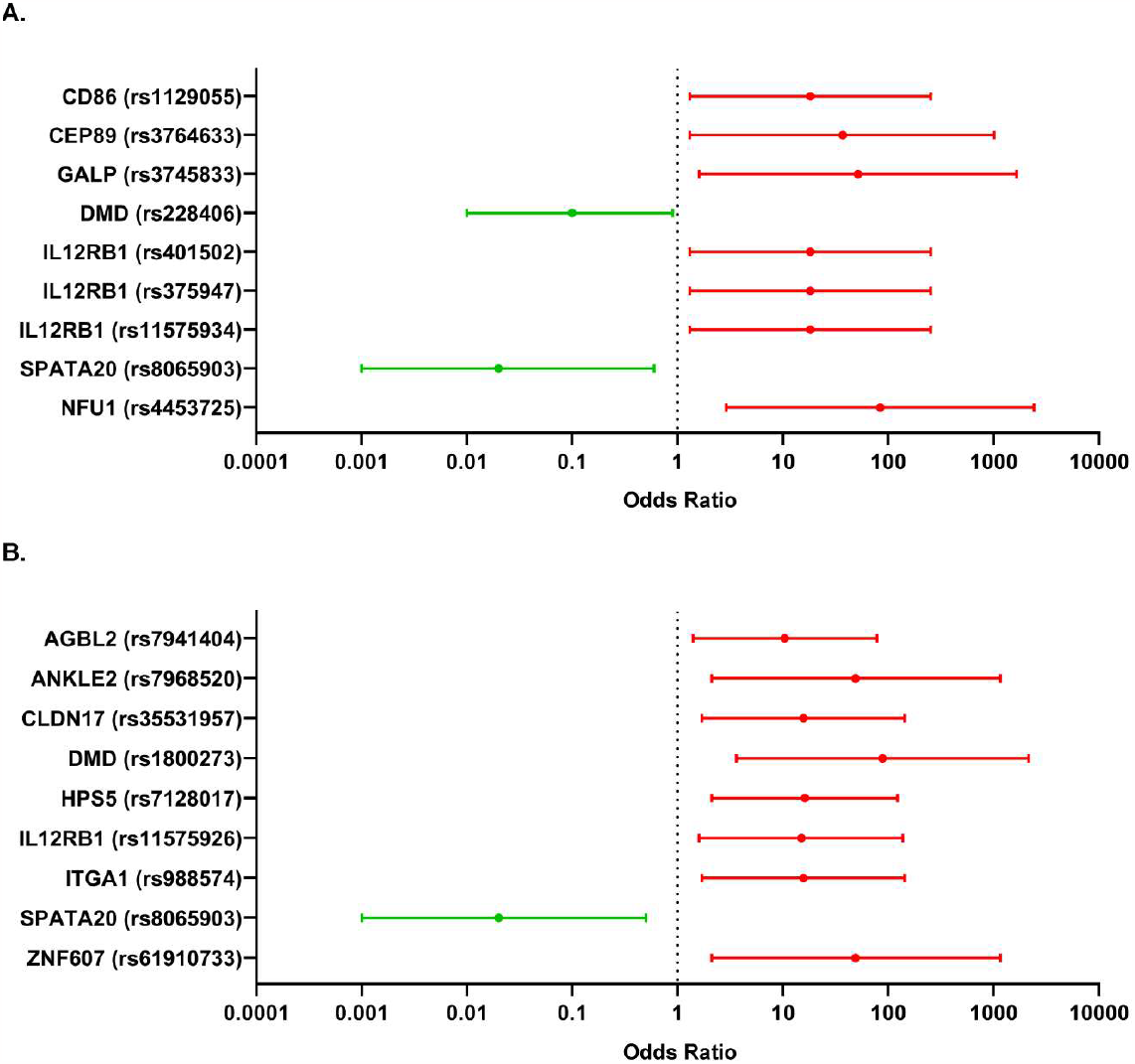
Forest plot showing the significant variants in candidate modifier genes associated to the FAST phenotype of FHHNC patients. **A**. Odds ratios and 95% confidence intervals were calculated by comparing the distribution of the alternative allele in homozygosis versus the reference allele in homozygosis. **B**. Odds ratios and 95% confidence intervals were calculated by comparing the distribution of the alternative allele in heterozygosis versus the reference allele in homozygosis.

### 3.4. Identified phenotype modifier variants are overrepresented in FHHNC patients with a faster renal phenotype progression compared to patients with moderate/mild phenotype and healthy individuals

In order to understand the functional implications of SNVs associated to renal progression in FHHNC, we retrieved expression quantitative trait loci (eQTL) analysis data for each significant variant in human kidney cortex tissues (n=73) from the GTEx database. Although none of the gene variants had an impact on gene expression in renal tissue (figure 4), strikingly, all the identified SNVs risk alleles for fast renal progression were strongly underrepresented in the GTEx kidney tissues (figure 4 and supplementary table 5).

**Figure 4.**
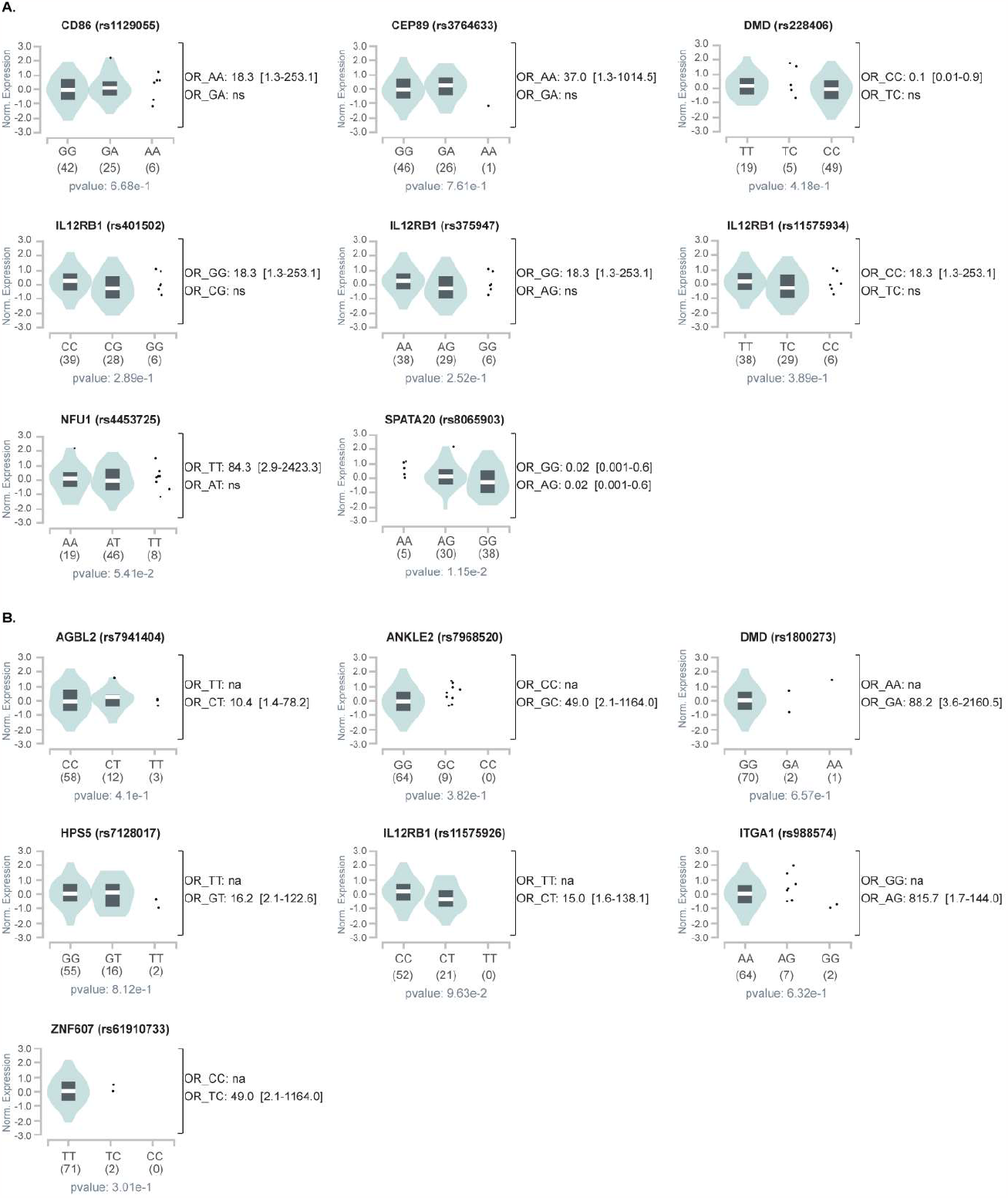
Violin plots of allele-specific cis-eQTLs according to genotypes for each significant variant in human kidney cortex tissues from the GTEx database. The teal region indicates the density distribution of the samples in each genotype. The allelic effect of variants on normalized gene expression levels are shown by boxplots within violin plots. The white line in the box plot (black) shows the median value of the expression of each genotype. The alleles and number of subjects analysed is shown under each genotype. **A**. Variants in homozygosis for the alternative allele associated to fast renal progression. **B**. Variants in heterozygosis associated to fast renal progression. The odds ratio and 95% confidence intervals calculated from table 2 are shown next to each plot for reference.

Conversely, we found that the FAST renal progression phenotype showed a higher proportion of patients harbouring the risk alleles than it would have been expected from the general healthy population here represented by the control tissues from GTEx (figure 5), and than what was also observed for the noFAST patients. Indeed, there were striking differences in the distribution of the three possible genotypes for each variant across the three groups and, particularly, comparing the FAST *versus* the noFAST phenotypes. For variants where the alternative allele in homozygosis was associated to the FAST phenotype, the proportion of subjects showing the alternative allele in homozygosis was significantly lower in the noFAST and GTEx groups (figure 5A). Conversely, variants rs228406 (*DMD*) and rs8065903 (*SPATA20*) for which the alternative allele in homozygosis was associated to the noFAST phenotype showed a higher proportion of subjects harbouring the reference allele in homozygosis in the FAST group (figure 5A). Additionally, heterozygotic variants associated to the FAST phenotype showed a significantly higher proportion of heterozygotic subjects in the FAST group than in the noFAST or GTEx groups (figure 5B).

**Figure 5.**
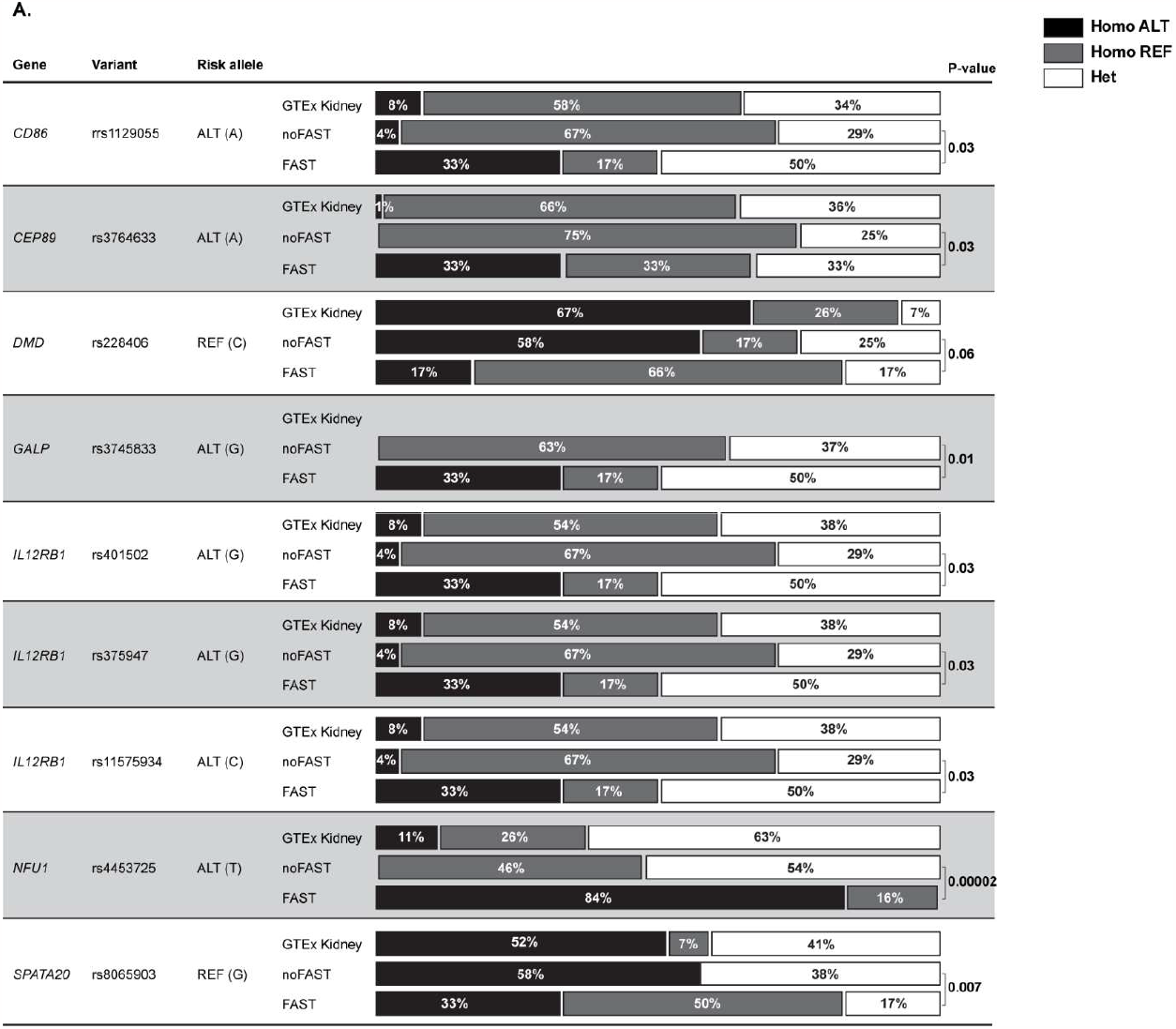
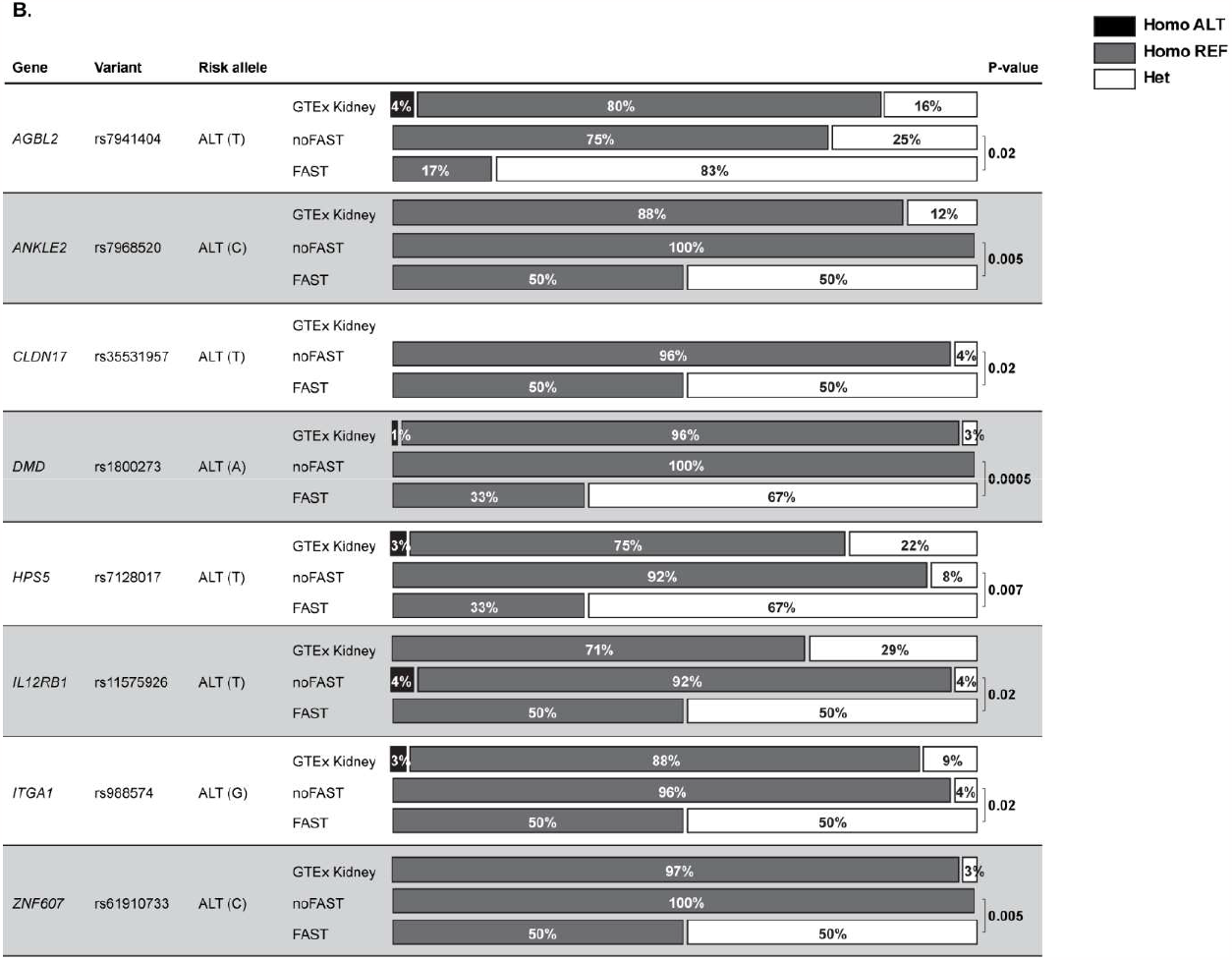
Distribution of the three possible genotypes for each candidate modifier gene variants across two FHHNC phenotypes and the control group from GTEx. **A**. Variants in homozygosis for the alternative allele associated to fast renal progression. **B**. Variants in heterozygosis associated to fast renal progression. Comparisons were performed by the Fisher’s exact test.

Next we aimed to identify whether the SNVs found showed a combined effect on the renal phenotype. Unfortunately, polygenic risk scores could not be determined as these SNVs are not annotated in any genome-wide study for FHHNC-related traits. However, 4 patients in the FAST group (67%) carried between 9 and 15 SNVs in the candidate modifier genes, while the maximum number of SNVs found in the noFAST group was 4 (supplementary table 6). Moreover, 9 noFAST patients (38%) did not carried any SNV and 10 patients (42%) carried only 1 or 2 SNVs in those candidate genes (supplementary table 7). Additionally, comparing the genetic background between siblings we found no substantial diferences in the number of prioritized SNVs carried between FHHNC patients and their healthy siblings (supplementary table 7). We had only one family with a FAST patient, a noFAST and a healthy sibling that were carrying 3 SNVs each and none were shared among the three of them (family 1 in supplementary table 7). The FAST patient from this family was the youngest to receive renal transplant with renal failure at 16 months old and the transplant performed one month later. This patient also shows the lowest number of prioritized SNVs in the FAST group suggesting that for this particular patient other factors may account for the fast renal progression. Additional intra-familial comparisons showed no substantial differences between siblings classified at the same phenotype (supplementary table 7).

## 4. DISCUSSION

The FHHNC clinical course is highly variable, whereas some individuals require kidney replacement therapy early in childhood, others maintain a more stable renal function into adulthood suffering with only mild to moderate renal disease^12^. Major research efforts in the field are focused on identifying novel mutations in the known causing genes, *CLDN16* and *CLDN19*, while potential genetic factors underlying phenotypic variability within patients have been ignored, likely due to the difficulties inherent to performing exome-wide association approaches in an ultra-rare disease. In the study presented here we describe the pipeline for WES analysis and candidate prioritization that has allowed us to identify for the first time potential phenotype modifier gene variants associated to a faster renal progression in FHHNC patients.

Our study was specifically designed to maximise the chances of identifying genes with significant associations in a small patient cohort, which benefits from the following key features. First, the unique cohort comprising 30 patients, representing almost the totality of FHHNC diagnosed subjects in Spain up to January 2021, with detailed clinical and genetic characterization. Second, the high genetic homogeneity of our cohort with 90% of patients harbouring *CLDN19* mutations and, from those, 70% harbouring the p.G20D founder mutation in homozygosis. Third, we used an extreme phenotype study design based on the assumption that risk variants would segregate with a more severe phenotype. This approach has been successfully applied to other diseases and has been proven to increase power, particularly when dealing with limited sample sizes as in rare diseases^14,15^. Therefore, we stratified our patients at the extremes of a quantitative trait as the annual decline on the estimated glomerular filtration rate as previously reported by our group^12^. And fourth, we applied a WES data analysis pipeline assessing mixed effects of variants using SKAT-O. SKAT-O is a unified approach that optimally combines the burden and non-burden sequence kernel association test (SKAT) and increases statistical power to detect associations, particularly when used in combination with extreme phenotypes analysis strategy^32^.

By using SKAT-O and comparing patients with a fast renal progression to patients with a moderate/mild renal progression, we obtained a high number of genes associated to the most severe renal phenotype. As expected, due to the limited number of samples analysed, none of those candidate genes resisted multiple testing correction. However, we implemented a prioritization strategy aimed to identify the genes that could be more relevant to FHHNC according to the state-of-the-art knowledge from the disease and thus limiting, at least in part, the possibility of selecting false positives. This strategy consisted on i) reducing our threshold for significance to P<0.01, ii) filtering the list for those genes expressed in renal tissue, and iii) further filtering the list selecting genes that had been previously associated to FHHNC key phenotypic traits by GWAS. Remarkably, overrepresentation analysis on prioritized candidates identified apical junction regulation and assembly, cell adhesion and immune system-related processes as the most significant terms associated to the modifier variants in the fast renal progression group which indicates that our analysis pipeline is selecting the candidates that are better aligned to the physiopathological hallmarks of the disease and, thus, most likely to have a true involvement in its clinical course. We acknowledge, however, that the applied extra filtering steps by tissue expression and GWAS annotation may be suspicious of introducing some bias in the enrichment analysis (terms related to kidney or to the traits used for GWAS-annotation would probably be favoured due to the pre-filtering). However, it should be noted that the overrepresentation analysis has been applied only as means to functionally annotate the selected genes and it did not have any influence on the candidate prioritization strategy.

Taking all the above considerations together, we took a further step to minimize the chances of selecting false positives and to prioritize gene variants associated to a higher risk of a faster renal progression by calculating the Fisher’s exact test and odds ratios. This additional analysis reduced the prioritized genes to 17 variants in 13 genes associated to a faster renal progression. Variants in these genes are annotated in the GWAS catalog associated to Ca^2+^ levels, eGFR, blood urea nitrogen levels, microalbuminuria, bone mineral density and high myopia, although the precise mechanisms underlying these associations have not been explored. Moreover, the variants found in our FHHNC cohort are different from those annotated in the GWAS catalog and their involvement in renal failure progression remains to be elucidated. Interestingly, among the genes conferring increased risk to faster renal progression, the biggest effect size corresponded to the homozygotic variant p.M25K in gene *NFU1. NFU1* is involved in iron–sulfur [Fe-S] cluster assembly, a highly complex system involved in several cellular processes including mitochondrial respiratory chain activity and various other enzymatic and regulatory functions^33-35^. Defects in this process have a severe impact on human health^33,36^ and, indeed, loss-of-function mutations in *NFU1* and other proteins involved in [Fe-S] cluster assembly have been described in multiple mitochondrial dysfunction syndromes (MMDS), a rare disease characterized by neurological regression, reduced motor control (dystonia) and pulmonary hypertension. Although almost all *NFU1* pathogenic mutations described so far are located in the Cterminal NifU [Fe-S] cluster binding domain^37^, the *NFU1* p.R21P mutation located in the N-domain, which is only 4 aa apart from p.M25K and affects the mitochondrial target signal^38^, has been described in the only MMDS patient showing renal tubular impairment^37,39^. Renal involvement has been also described for MMDS patients harbouring mutations in *BOLA3*^37^, a protein that cooperates with *NFU1* in the last steps of maturation and transfer of [4Fe-4S] clusters into target proteins, further supporting the role of [Fe-S] cluster assembly defects on kidney disease. Our results suggest that [Fe-S] cluster homeostasis may be imbalanced in renal tubular cells from FHHNC patients with a more severe renal phenotype contributing, therefore, to the aggravation of the high cellular stress already produced by the *CLDN19* mutation. The potential involvement of the p.M25K variant in *NFU1* in FHHNC renal failure progression is further supported by the fact that this variant is considered a common variant with a MAF of 0.39 according to the gnomAD browser. Therefore, its potential impact on cellular homeostasis is likely to be easily accommodated in healthy individuals particularly when the variant is present in heterozygosis, as it happens in the majority of cases genotyped. The homozygotic variant represents only around 8% of cases in the general population and around 20% of the total of cases harbouring the alternative allele for this variant. In concordance, the GTEx data showed that the p.M25K *NFU1* variant was present in homozygosis in only 11% of the genotyped renal tissues. Remarkably, none of the FHHNC with a moderate/mild progression had this variant in homozygosis, while it was present in 5 out of 6 patients with a fast renal progression. This striking accumulation of the *NFU1* risk variant in our FHHNC patients with a more severe phenotype was a common observation for almost all the other variants prioritized in our study. Of particular interest are the variants found in the genes encoding dystrophin (*DMD*) and Hermansky-Pudlak Syndrome protein 5 (*HPS5*), as mutations in both genes have been associated to rare diseases accompanied with compromised renal function^40^. Indeed, progressive muscle degeneration in Duchene’s muscular dystrophy, caused by mutations in *DMD*, seems to be associated to subclinical kidney injury and the subsequent development of chronic kidney disease in paediatric patients^41^. Furthermore, and although insufficiently explored, neuromuscular impairment has been suggested to be part of the clinical spectrum associated with *CLDN19* mutations as intolerance to muscular exercise that persist after kidney transplant and normalization of magnesium levels has been described in a few patients^42^. On the other hand, the HPS5 protein is highly expressed in human renal tubules according to the Human Protein Atlas and HPS5 knockdown resulted in a severe renal phenotype in a zebrafish model through a mechanism that involves intracellular accumulation of cell debris due to lysosomal impairment^40^. In addition, a variant in another member of the claudin family of proteins, *CLDN17*, is also associated to a more aggressive phenotype in our FHHNC cohort. Data on *CLDN17* is scarce in the scientific literature and the GWAS catalog filter applied in our prioritization pipeline failed to identify this gene as a candidate for further analysis. However, claudins play a critical role on FHHNC and CLDN17 protein, but not RNA, has been described to be present in mouse and human proximal tubules^6^, therefore we decided to include it in our prioritized SKAT-O gene list. A crucial role for *CLDN17* in renal function via the regulation of serum electrolytes and tissue reactive oxygen species levels has been recently reported^43^. Although promising, the functional impact on the FHHNC specific variants in *NFU1, DMD, HPS5* and *CLDN17* and how they contribute to faster renal progression towards kidney failure remains to be characterized. We acknowledge that although replication studies in an independent cohort is also highly recommended, it would be extremely difficult to gather enough patients in a reasonable time frame to do so. In addition, functional data from the specific variants identified in patients with the most severe outcome would be desirable to understand their involvement in renal progression in our patients. However, it is important to remark that even gathering information regarding the impact of the genetic variants on their respective protein function in cell and/or animal models, there would be still a huge gap to explain their contribution to such a complex phenotype as in FHHNC. Furthermore, since these variants have not been described to be pathogenic per se, in light of our results we can speculate that they interact together as part of the genetic context of patients with a more severe renal phenotype. How these variants are combined to interact with the FHHNC causative *CLDN19* mutation resulting in a particular renal phenotype is far more difficult to approach. Appropriate models for this disease, which arises from the thick ascending limb of Henle’s loop of the nephron, simply do not exist and the knock-in mouse model carrying the *CLDN19* p.G20D causative mutation has not been generated either. Moreover, it is not ethical to perform renal biopsies in these patients so we cannot even have access to patient’s tissue and had to rely on data from the GTEx database as representative from the general population.

In summary, we have applied a pipeline specifically tailored for an ultra-rare renal disease that has allowed us to find for the first time several gene variants associated with a more aggressive phenotype. Particularly, the most promising data comes from variants that may affect the functional stability of cell organelles like the mitochondria and lysosomes. The orchestrated action of these organelles is key for cellular homeostasis and survival under pathological stress conditions^44^ and may explain the variability of the clinical phenotype in FHHNC in which the endoplasmic reticulum (ER) is likely to be under high stress due to the reported retention of *CLDN19* p.G20D mutant protein^5^. The fact that our candidate modifier genes are also associated with other diseases involving signs of kidney disease also brings up the interesting possibility that their involvement in renal failure progression may not be specific for FHHNC. Potential prognostic and therapeutic tools developed for these targets could, thus, be applicable to a wider spectrum of renal and extra-renal diseases. Finally, our approach can easily be customized to identify and study modifier genes for other forms of renal rare diseases and can contribute to strengthen the interpretation of personalized exome sequencing approaches providing, therefore, a valuable framework for the identification and understanding of phenotypic variability and disease risk.

## Data Availability

Availability of data and materials: De-identified data will be made available upon request from qualified investigators studying the molecular basis of renal rare disorders. Datasets can be obtained via the corresponding author.

## Acknowledgements

We would like thank the FHHNC patient advocacy group HIPOFAM (http://hipofam.org), patients and families for their valuable support and contribution to our research activity. We also thank Dr. Xavier de la Cruz for helpful discussions and advise. The Genotype-Tissue Expression (GTEx) Project was supported by the Common Fund of the Office of the Director of the National Institutes of Health, and by NCI, NHGRI, NHLBI, NIDA, NIMH, and NINDS.

## Author Contributions

MV-P: sample collection and processing, data analysis; MD, JT: sample collection and processing; JM, RT, MF, AS: data analysis; GC-R: review-editing; GA: conceptualization, funding acquisition, patient recruitment, clinical data analysis and supervision, review-editing; AM: conceptualization, funding acquisition, methodology, supervision, review-editing; CM: conceptualization, methodology, data analysis, writing original draft, review-editing. All authors have read and agreed to the published version of the manuscript.

## Availability of data and materials

De-identified data will be made available upon request from qualified investigators studying the molecular basis of renal rare disorders. Datasets can be obtained via the corresponding author.

