## Supplemental data for "IDENTIFICATION OF MODIFIER GENE VARIANTS OVERREPRESENTED IN FAMILIAL HYPOMAGNESEMIA WITH HYPERCALCIURIA AND NEPHROCALCINOSIS PATIENTS WITH A MORE AGGRESSIVE RENAL PHENOTYPE"

Vall-Palomar, *et al*.

### SUPPLEMENTARY DATA

#### Supplementary table 1. Number of patients distributed in each group.

| Genetics | | Renal Phenotype (according to decline in eGFR) | | | Visual impairment | |
|---|---|---|---|---|---|---|
| | | Fast | Moderate | Slow | YES | NO |
| **CLDN16** | | 0 | 1 | 2 | 0 | 3 |
| **CLDN19** | p.G20D homozygous | 4 | 12 | 4 | 17 | 3 |
| | p.G20D heterozygous + other variants | 2 | 4 | 1 | 3 | 2 |

CLDN16: claudin 16; CLDN19: claudin 19; eGFR: estimated glomerular filtration rate

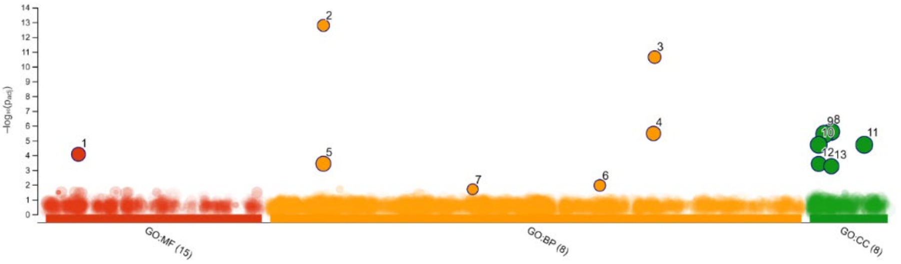


| ID | Source | Term ID | Term Name | $p_{adj}$ (query_1) |
|---|---|---|---|---|
| 1 | GO:MF | GO:0005509 | calcium ion binding | $8.545 \times 10^{-5}$ |
| 2 | GO:BP | GO:0007156 | homophilic cell adhesion via plasma membrane a... | $1.626 \times 10^{-13}$ |
| 3 | GO:BP | GO:0098742 | cell-cell adhesion via plasma-membrane adhesion ... | $2.268 \times 10^{-11}$ |
| 4 | GO:BP | GO:0098609 | cell-cell adhesion | $3.334 \times 10^{-6}$ |
| 5 | GO:BP | GO:0007155 | cell adhesion | $3.714 \times 10^{-4}$ |
| 6 | GO:BP | GO:0070555 | response to interleukin-1 | $1.098 \times 10^{-2}$ |
| 7 | GO:BP | GO:0042246 | tissue regeneration | $1.989 \times 10^{-2}$ |
| 8 | GO:CC | GO:0031224 | intrinsic component of membrane | $2.566 \times 10^{-6}$ |
| 9 | GO:CC | GO:0016020 | membrane | $3.665 \times 10^{-6}$ |
| 10 | GO:CC | GO:0005886 | plasma membrane | $1.951 \times 10^{-5}$ |
| 11 | GO:CC | GO:0071944 | cell periphery | $1.951 \times 10^{-5}$ |
| 12 | GO:CC | GO:0005887 | integral component of plasma membrane | $3.835 \times 10^{-4}$ |
| 13 | GO:CC | GO:0031226 | intrinsic component of plasma membrane | $5.525 \times 10^{-4}$ |

**Supplementary figure 1. Graphic representation of g:Profiler functional analysis of the candidate modifier genes obtained by SKAT-O.** The most significant results for Gene Ontology (GO) are shown. GO molecular function (GO:MF); GO biological process (GO:BP).

**Supplementary table 2. Phenotypic traits related to FHHNC and matched SKAT-O genes to GWAS catalog associated variants.**

| Trait | Associated variants found in GWAS catalog | Matched SKATO-genes |
|---|---|---|
| glomerular filtration rate | 3729 | 16 |
| chronic kidney disease | 418 | 7 |
| nephritis | 161 | 0 |
| nephrolithiasis | 43 | 0 |
| nephrocalcinosis | 0 | 0 |
| susceptibility to urinary tract infection measurement | 61 | 0 |
| renal system measurement | 5000 | 20 |
| proteinuria | 207 | 2 |
| urinary metabolite measurement | 1358 | 4 |
| magnesium measurement | 64 | 0 |
| calcium measurement | 778 | 7 |
| parathyroid hormone measurement | 37 | 0 |
| vitamin D measurement | 713 | 3 |
| bone density | 5000 | 8 |
| macular degeneration | 302 | 5 |
| myopia | 837 | 4 |
| retinopathy | 553 | 3 |

**Supplementary table 3. Significant single nucleotide variants associated to the fast renal progression phenotype.**

| | Gene Name | Transcript ID | Variant ID | RA | AA | HGVSc | HGVSp | Most severe consequence | MAF | Fisher test P-value | Odds Ratio [95% CI] |
|---|---|---|---|---|---|---|---|---|---|---|---|
| **Homozigous alternative allele** | *CD86* | ENST00000330540 | rs1129055 | G | A | c.928G>A | p.Ala310Thr | missense_variant | 0.30 | 0.03 | 18.3 [1.3-253.1] |
| | *CEP89* | ENST00000305768 | rs3764633 | G | A | c.580C>T | p.Arg194Trp | missense_variant | 0.18 | 0.03 | 37.0 [1.3-1014.5] |
| | *DMD* | ENST00000357033 | rs228406 | T | C | c.2645A>G | p.Asp882Gly | missense_variant | 0.72 | 0.06 | 0.1 [0.01-0.9] |
| | *GALP* | ENST00000357330 | rs3745833 | C | G | c.216C>G | p.Ile72Met | missense_variant | 0.34 | 0.01 | 51.7 [1.6-1657.5] |
| | *IL12RB1* | ENST00000593993 | rs401502 | C | G | c.1132G>C | p.Gly378Arg | missense_variant | 0.28 | 0.03 | 18.3 [1.3-253.1] |
| | | | rs375947 | A | G | c.1094T>C | p.Met365Thr | missense_variant | 0.29 | 0.03 | 18.3 [1.3-253.1] |
| | | | rs11575934 | T | C | c.641A>G | p.Gln214Arg | missense_variant | 0.28 | 0.03 | 18.3 [1.3-253.1] |
| | *NFU1* | ENST00000410022 | rs4453725 | A | T | c.74T>A | p.Met25Lys | missense / start loss | 0.39 | 0.00002 | 84.3 [2.9-2423.3] |
| | *SPATA20* | ENST00000006658 | rs8065903 | A | G | c.1874A>G | p.Lys625Arg | missense_variant | 0.79 | 0.01 | 0.02 [0.001-0.6] |
| **Heterozigous** | *AGBL2* | ENST00000357610 | rs7941404 | C | T | c.1046G>A | p.Arg349His | missense_variant | 0.09 | 0.02 | 10.4 [1.4-78.2] |
| | *ANKLE2* | ENST00000357997 | rs7968520 | G | C | c.442C>G | p.Gln148Glu | missense_variant | 0.08 | 0.005 | 49.0 [2.1-1164.0] |
| | *CLDN17* | ENST00000286808 | rs35531957 | C | T | c.244G>A | p.Ala82Thr | missense_variant | 0.10 | 0.02 | 15.7 [1.7-144.0] |
| | *DMD* | ENST00000357033 | rs1800273 | G | A | c.6463C>T | p.Arg2155Trp | missense_variant | 0.03 | 0.0005 | 88.2 [3.6-2160.5] |
| | *HPS5* | ENST00000349215 | rs7128017 | G | T | c.1249C>A | p.Leu417Met | missense_variant | 0.14 | 0.01 | 16.2 [2.1-122.6] |
| | *IL12RB1* | ENST00000593993 | rs11575926 | C | T | c.467G>A | p.Arg156His | missense_variant | 0.13 | 0.02 | 15.0 [1.6-138.1] |
| | *ITGA1* | ENST00000282588 | rs988574 | A | G | c.3323A>G | p.Glu1108Gly | missense_variant | 0.06 | 0.02 | 15.7 [1.7-144.0] |
| | *SPATA20* | ENST00000006658 | rs8065903 | A | G | c.1874A>G | p.Lys625Arg | missense_variant | 0.79 | 0.01 | 0.02 [0.001-0.6] |
| | *ZNF607* | ENST00000355202 | rs61910733 | T | C | c.1477A>G | p.Thr493Ala | missense_variant | 0.01 | 0.005 | 49.0 [2.1-1164.0] |

HGVSc: nomenclature for the Human Genome Variant Society, coding DNA reference sequence; HGVSp: nomenclature for the Human Genome Variant Society, protein reference sequence; MAF: minor allele frequency (from gnomAD); CI: confidence interval; RA: reference allele; AA: alternative allele.

### Supplementary table 4. Prioritized phenotype modifier candidates and associated cluster and gene ontology biological process (GO-BP) from summORA analysis.

| Prioritized SKAT-O genes | Cluster | GO-BP term | P-value | P.adjust |
|---|---|---|---|---|
| CD86 | 3 | lymphotoxin A production | 0.008 | 0.18 |
| | | CD40 signaling pathway | 0.04 | 0.22 |
| | 4 | positive regulation of T-helper cell differentiation | 0.001 | 0.16 |
| | | positive regulation of T cell differentiation | 0.002 | 0.16 |
| | | positive regulation of lymphocyte differentiation | 0.003 | 0.18 |
| | | positive regulation of CD4-positive, alpha-beta T cell differentiation | 0.003 | 0.18 |
| | | regulation of T-helper cell differentiation | 0.004 | 0.18 |
| | | positive regulation of CD4-positive, alpha-beta T cell activation | 0.004 | 0.18 |
| | | regulation of T cell differentiation | 0.007 | 0.18 |
| | | regulation of CD4-positive, alpha-beta T cell differentiation | 0.007 | 0.18 |
| | | positive regulation of alpha-beta T cell differentiation | 0.007 | 0.18 |
| | | positive regulation of leukocyte differentiation | 0.008 | 0.18 |
| | | positive regulation of hemopoiesis | 0.008 | 0.18 |
| | | regulation of T cell proliferation | 0.01 | 0.20 |
| | | regulation of lymphocyte differentiation | 0.01 | 0.20 |
| | | regulation of T cell activation | 0.01 | 0.20 |
| | | T-helper cell differentiation | 0.01 | 0.20 |
| | | regulation of CD4-positive, alpha-beta T cell activation | 0.01 | 0.20 |
| | | CD4-positive, alpha-beta T cell differentiation involved in immune response | 0.01 | 0.20 |
| | | regulation of alpha-beta T cell differentiation | 0.01 | 0.20 |
| | | alpha-beta T cell activation involved in immune response | 0.01 | 0.20 |
| | | alpha-beta T cell differentiation involved in immune response | 0.01 | 0.20 |
| | | negative regulation of T cell proliferation | 0.01 | 0.20 |
| | | positive regulation of alpha-beta T cell activation | 0.01 | 0.20 |
| | | T cell proliferation | 0.02 | 0.20 |
| | | T cell differentiation involved in immune response | 0.02 | 0.20 |
| | | positive regulation of T-helper 2 cell differentiation | 0.02 | 0.20 |
| | | CD4-positive, alpha-beta T cell differentiation | 0.02 | 0.20 |
| | | positive regulation of T cell activation | 0.02 | 0.20 |
| | | negative regulation of lymphocyte proliferation | 0.02 | 0.20 |
| | | regulation of lymphocyte proliferation | 0.02 | 0.21 |
| | | regulation of T-helper 2 cell differentiation | 0.03 | 0.21 |
| | | CD4-positive, alpha-beta T cell activation | 0.03 | 0.21 |
| | | positive regulation of T cell proliferation | 0.03 | 0.21 |
| | | T cell differentiation | 0.03 | 0.21 |
| | | regulation of alpha-beta T cell activation | 0.03 | 0.21 |
| | | alpha-beta T cell differentiation | 0.03 | 0.21 |
| | | T cell activation involved in immune response | 0.04 | 0.22 |
| | | regulation of leukocyte differentiation | 0.04 | 0.22 |
| | | T-helper 2 cell differentiation | 0.04 | 0.22 |
| | | lymphocyte proliferation | 0.04 | 0.22 |
| | | negative regulation of T cell activation | 0.04 | 0.22 |
| | | positive regulation of type 2 immune response | 0.04 | 0.22 |
| | 7 | regulation of lymphotoxin A production | 0.008 | 0.18 |
| | | positive regulation of lymphotoxin A production | 0.008 | 0.18 |
| | | activation of protein kinase C activity | 0.01 | 0.20 |
| | 9 | regulation of leukocyte cell-cell adhesion | 0.01 | 0.20 |
| | | leukocyte cell-cell adhesion | 0.02 | 0.20 |
| | | positive regulation of leukocyte cell-cell adhesion | 0.02 | 0.21 |
| | | regulation of cell-cell adhesion | 0.03 | 0.21 |
| | | positive regulation of cell-cell adhesion | 0.04 | 0.22 |
| | 10 | regulation of lipase activity | 0.02 | 0.21 |
| | 12 | negative regulation of mononuclear cell proliferation | 0.02 | 0.20 |
| | | regulation of mononuclear cell proliferation | 0.02 | 0.21 |
| | | negative regulation of leukocyte proliferation | 0.02 | 0.21 |
| | | regulation of leukocyte proliferation | 0.03 | 0.21 |
| | | mononuclear cell proliferation | 0.04 | 0.22 |
| | 3 | regulation of skeletal muscle contraction by calcium ion signaling | 0.01 | 0.20 |
| | | regulation of skeletal muscle contraction | 0.04 | 0.22 |
| | 6 | regulation of skeletal muscle contraction by regulation of release of sequestered calcium ion | 0.01 | 0.20 |
| | | sodium ion transmembrane transport | 0.01 | 0.20 |
| | | sodium ion transport | 0.03 | 0.21 |
| | | positive regulation of sodium ion transmembrane transporter activity | 0.04 | 0.22 |
| | 7 | negative regulation of peptidyl-serine phosphorylation | 0.003 | 0.18 |
| | | peptidyl-cysteine modification | 0.007 | 0.18 |
| | | negative regulation of peptidyl-cysteine S-nitrosylation | 0.02 | 0.20 |
| | | regulation of peptidyl-cysteine S-nitrosylation | 0.03 | 0.21 |
| | | protein nitrosylation | 0.04 | 0.22 |
| | | peptidyl-cysteine S-nitrosylation | 0.04 | 0.22 |
| GALP | 3 | neuropeptide signaling pathway | 0.03 | 0.21 |

| | | | | |
|---|---|---|---|---|
| IL12RB1 | 3 | positive regulation of defense response to virus by host | 0.003 | 0.18 |
| | | regulation of defense response to virus by host | 0.005 | 0.18 |
| | | interleukin-23-mediated signaling pathway | 0.008 | 0.18 |
| | | interleukin-12-mediated signaling pathway | 0.01 | 0.20 |
| | | cellular response to interleukin-12 | 0.02 | 0.20 |
| | | regulation of defense response to virus | 0.02 | 0.20 |
| | | response to interleukin-12 | 0.02 | 0.20 |
| | | immunological memory formation process | 0.04 | 0.22 |
| | | immunological memory process | 0.04 | 0.22 |
| | 4 | positive regulation of T-helper cell differentiation | 0.001 | 0.16 |
| | | positive regulation of T cell differentiation | 0.002 | 0.16 |
| | | positive regulation of lymphocyte differentiation | 0.003 | 0.18 |
| | | positive regulation of CD4-positive, alpha-beta T cell differentiation | 0.003 | 0.18 |
| | | regulation of T-helper cell differentiation | 0.004 | 0.18 |
| | | positive regulation of CD4-positive, alpha-beta T cell activation | 0.004 | 0.18 |
| | | regulation of T cell differentiation | 0.007 | 0.18 |
| | | regulation of CD4-positive, alpha-beta T cell differentiation | 0.007 | 0.18 |
| | | positive regulation of alpha-beta T cell differentiation | 0.007 | 0.18 |
| | | positive regulation of leukocyte differentiation | 0.008 | 0.18 |
| | | positive regulation of hemopoiesis | 0.008 | 0.18 |
| | | regulation of T cell proliferation | 0.01 | 0.20 |
| | | positive regulation of T-helper 17 cell lineage commitment | 0.01 | 0.20 |
| | | regulation of lymphocyte differentiation | 0.01 | 0.20 |
| | | regulation of T cell activation | 0.01 | 0.20 |
| | | T-helper cell differentiation | 0.01 | 0.20 |
| | | regulation of CD4-positive, alpha-beta T cell activation | 0.01 | 0.20 |
| | | CD4-positive, alpha-beta T cell differentiation involved in immune response | 0.01 | 0.20 |
| | | regulation of alpha-beta T cell differentiation | 0.01 | 0.20 |
| | | alpha-beta T cell activation involved in immune response | 0.01 | 0.20 |
| | | alpha-beta T cell differentiation involved in immune response | 0.01 | 0.20 |
| | | positive regulation of alpha-beta T cell activation | 0.01 | 0.20 |
| | | T cell proliferation | 0.02 | 0.20 |
| | | regulation of T-helper 17 cell lineage commitment | 0.02 | 0.20 |
| | | T cell differentiation involved in immune response | 0.02 | 0.20 |
| | | CD4-positive, alpha-beta T cell differentiation | 0.02 | 0.20 |
| | | positive regulation of T cell activation | 0.02 | 0.20 |
| | | positive regulation of T-helper 17 cell differentiation | 0.02 | 0.20 |
| | | regulation of lymphocyte proliferation | 0.02 | 0.21 |
| | | positive regulation of memory T cell differentiation | 0.02 | 0.21 |
| | | CD4-positive, alpha-beta T cell activation | 0.03 | 0.21 |
| | | positive regulation of T cell proliferation | 0.03 | 0.21 |
| | | T cell differentiation | 0.03 | 0.21 |
| | | regulation of alpha-beta T cell activation | 0.03 | 0.21 |
| | | positive regulation of cell fate commitment | 0.03 | 0.21 |
| | | regulation of memory T cell differentiation | 0.03 | 0.21 |
| | | alpha-beta T cell differentiation | 0.03 | 0.21 |
| | | memory T cell differentiation | 0.03 | 0.21 |
| | | T-helper 17 cell lineage commitment | 0.03 | 0.21 |
| | | positive regulation of T-helper 17 type immune response | 0.04 | 0.22 |
| | | T cell activation involved in immune response | 0.04 | 0.22 |
| | | regulation of leukocyte differentiation | 0.04 | 0.22 |
| | | lymphocyte proliferation | 0.04 | 0.22 |
| | | T-helper cell lineage commitment | 0.04 | 0.22 |
| | | positive regulation of T-helper 1 type immune response | 0.04 | 0.23 |
| | | regulation of T-helper 17 cell differentiation | 0.04 | 0.23 |
| | 9 | regulation of leukocyte cell-cell adhesion | 0.01 | 0.20 |
| | | leukocyte cell-cell adhesion | 0.02 | 0.20 |
| | | positive regulation of leukocyte cell-cell adhesion | 0.02 | 0.21 |
| | | regulation of cell-cell adhesion | 0.03 | 0.21 |
| | | positive regulation of cell-cell adhesion | 0.04 | 0.22 |
| | 12 | regulation of mononuclear cell proliferation | 0.02 | 0.21 |
| | | regulation of leukocyte proliferation | 0.03 | 0.21 |
| | | mononuclear cell proliferation | 0.04 | 0.22 |
| NFU1 | 7 | protein maturation by iron-sulfur cluster transfer | 0.04 | 0.22 |
| AGBL2 | 7 | protein side chain deglutamylation | 0.02 | 0.20 |
| | | protein deglutamylation | 0.02 | 0.20 |
| ANKLE2 | 7 | positive regulation of protein dephosphorylation | 0.006 | 0.18 |
| | | positive regulation of dephosphorylation | 0.01 | 0.20 |
| | | regulation of protein dephosphorylation | 0.02 | 0.21 |
| | | regulation of dephosphorylation | 0.04 | 0.23 |
| | 8 | mitotic nuclear membrane reassembly | 0.02 | 0.21 |
| | | mitotic nuclear membrane organization | 0.02 | 0.21 |
| CLDN17 | 8 | apical junction assembly | 0.00004 | 0.02 |
| | | cell-cell junction assembly | 0.0006 | 0.11 |
| | | bicellular tight junction assembly | 0.0007 | 0.12 |
| | | tight junction assembly | 0.0009 | 0.14 |
| | | tight junction organization | 0.001 | 0.15 |
| | | cell-cell junction organization | 0.002 | 0.16 |
| | | cell junction assembly | 0.02 | 0.21 |
| | 9 | calcium-independent cell-cell adhesion via plasma membrane cell-adhesion molecules | 0.002 | 0.16 |
| ITGA1 | 7 | positive regulation of protein dephosphorylation | 0.006 | 0.18 |
| | | positive regulation of dephosphorylation | 0.01 | 0.20 |
| | | regulation of protein dephosphorylation | 0.02 | 0.21 |
| | | regulation of dephosphorylation | 0.04 | 0.23 |

**Supplementary table 5. Distribution of genotypes for each variant across the FHHNC phenotypes and the GTEx control group.**

| | Gene Name | Variant ID | Risk allele | Genotype | FAST (n) | NoFAST (n) | GTEx (n) |
|---|---|---|---|---|---|---|---|
| Homozigous alternative allele | *CD86* | rs1129055 | ALT (A) | HomALT | 2 | 1 | 6 |
| | | | | HomREF | 1 | 16 | 42 |
| | | | | Het | 3 | 7 | 25 |
| | *CEP89* | rs3764633 | ALT (A) | HomALT | 2 | 0 | 1 |
| | | | | HomREF | 2 | 18 | 46 |
| | | | | Het | 2 | 6 | 26 |
| | *DMD* | rs228406 | REF (C) | HomALT | 1 | 14 | 49 |
| | | | | HomREF | 4 | 4 | 19 |
| | | | | Het | 1 | 6 | 5 |
| | *GALP* | rs3745833 | ALT (G) | HomALT | 2 | 0 | na |
| | | | | HomREF | 1 | 15 | na |
| | | | | Het | 3 | 9 | na |
| | *IL12RB1* | rs401502 | ALT (G) | HomALT | 2 | 1 | 6 |
| | | | | HomREF | 1 | 16 | 39 |
| | | | | Het | 3 | 7 | 28 |
| | | rs375947 | ALT (G) | HomALT | 2 | 1 | 6 |
| | | | | HomREF | 1 | 16 | 38 |
| | | | | Het | 3 | 7 | 29 |
| | | rs11575934 | ALT (C) | HomALT | 2 | 1 | 6 |
| | | | | HomREF | 1 | 16 | 38 |
| | | | | Het | 3 | 7 | 29 |
| | *SPATA20* | rs8065903 | REF (G) | HomALT | 2 | 14 | 8 |
| | | | | HomREF | 3 | 0 | 19 |
| | | | | Het | 1 | 10 | 46 |
| | *NFU1* | rs4453725 | ALT (T) | HomALT | 5 | 0 | 38 |
| | | | | HomREF | 1 | 11 | 5 |
| | | | | Het | 0 | 13 | 30 |
| Heterozigous | *AGBL2* | rs7941404 | ALT (T) | HomALT | 0 | 0 | 3 |
| | | | | HomREF | 1 | 18 | 58 |
| | | | | Het | 5 | 6 | 12 |
| | *ANKLE2* | rs7968520 | ALT (C) | HomALT | 0 | 0 | 0 |
| | | | | HomREF | 3 | 24 | 64 |
| | | | | Het | 3 | 0 | 9 |
| | *CLDN17* | rs35531957 | ALT (T) | HomALT | 0 | 0 | na |
| | | | | HomREF | 3 | 23 | na |
| | | | | Het | 3 | 1 | na |
| | *DMD* | rs1800273 | ALT (A) | HomALT | 0 | 0 | 1 |
| | | | | HomREF | 2 | 24 | 70 |
| | | | | Het | 4 | 0 | 2 |
| | *HPS5* | rs7128017 | ALT (T) | HomALT | 0 | 0 | 2 |
| | | | | HomREF | 2 | 22 | 55 |
| | | | | Het | 4 | 2 | 16 |
| | *IL12RB1* | rs11575926 | ALT (T) | HomALT | 0 | 1 | 0 |
| | | | | HomREF | 3 | 22 | 52 |
| | | | | Het | 3 | 1 | 21 |
| | *ITGA1* | rs988574 | ALT (G) | HomALT | 0 | 0 | 2 |
| | | | | HomREF | 3 | 23 | 64 |
| | | | | Het | 3 | 1 | 7 |
| | *SPATA20* | rs8065903 | REF (G) | HomALT | 2 | 14 | 8 |
| | | | | HomREF | 3 | 0 | 19 |
| | | | | Het | 1 | 10 | 46 |
| | *ZNF607* | rs61910733 | ALT (C) | HomALT | 0 | 0 | 0 |
| | | | | HomREF | 3 | 24 | 71 |
| | | | | Het | 3 | 0 | 2 |

**Supplementary table 6. Presence of risk alleles for the prioritized candidate SNVs showing aggregation in the FHHNC FAST renal progression phenotype.**

| Gene ID | Variant ID | FAST phenotype | | | | | | noFAST renal phenotype | | | | | | | | | | | | | | | | | | | | | | | |
|---|---|---|---|---|---|---|---|---|---|---|---|---|---|---|---|---|---|---|---|---|---|---|---|---|---|---|---|---|---|---|---|
| | | | | | | | | Patients with kidney transplant | | | | | | | Patients with native kidney | | | | | | | | | | | | | | | | |
| CD86 | rs1129055 | | | | | | | | | | | | | | | | | | | | | | | | | | | | | | |
| CEP89 | rs3764633 | | | | | | | | | | | | | | | | | | | | | | | | | | | | | | |
| DMD | rs228406 | | | | | | | | | | | | | | | | | | | | | | | | | | | | | | |
| GALP | rs3745833 | | | | | | | | | | | | | | | | | | | | | | | | | | | | | | |
| IL12RB1 | rs401502 | | | | | | | | | | | | | | | | | | | | | | | | | | | | | | |
| IL12RB1 | rs375947 | | | | | | | | | | | | | | | | | | | | | | | | | | | | | | |
| IL12RB1 | rs11575934 | | | | | | | | | | | | | | | | | | | | | | | | | | | | | | |
| NFU1 | rs4453725 | | | | | | | | | | | | | | | | | | | | | | | | | | | | | | |
| SPATA20 | rs8065903 | | | | | | | | | | | | | | | | | | | | | | | | | | | | | | |
| AGBL2 | rs7941404 | | | | | | | | | | | | | | | | | | | | | | | | | | | | | | |
| ANKLE2 | rs7968520 | | | | | | | | | | | | | | | | | | | | | | | | | | | | | | |
| CLDN17 | rs35531957 | | | | | | | | | | | | | | | | | | | | | | | | | | | | | | |
| DMD | rs1800273 | | | | | | | | | | | | | | | | | | | | | | | | | | | | | | |
| HPS5 | rs7128017 | | | | | | | | | | | | | | | | | | | | | | | | | | | | | | |
| IL12RB1 | rs11575926 | | | | | | | | | | | | | | | | | | | | | | | | | | | | | | |
| ITGA1 | rs988574 | | | | | | | | | | | | | | | | | | | | | | | | | | | | | | |
| ZNF607 | rs61910733 | | | | | | | | | | | | | | | | | | | | | | | | | | | | | | |
| % Risk alleles | | 53% | 88% | 35% | 18% | 65% | 53% | 12% | 12% | 0% | 6% | 24% | 0% | 6% | 6% | 0% | 0% | 6% | 0% | 18% | 18% | 18% | 6% | 6% | 0% | 12% | 0% | 18% | 6% | 0% | 0% |
| % Non-risk alleles | | 47% | 12% | 65% | 82% | 35% | 47% | 88% | 88% | 100% | 94% | 76% | 100% | 94% | 94% | 100% | 100% | 94% | 100% | 82% | 82% | 82% | 94% | 94% | 100% | 88% | 100% | 82% | 94% | 100% | 100% |
| sum of risk alleles | | 9 | 15 | 6 | 3 | 11 | 9 | 2 | 2 | 0 | 1 | 4 | 0 | 1 | 1 | 0 | 0 | 1 | 0 | 3 | 3 | 3 | 1 | 1 | 0 | 2 | 0 | 3 | 1 | 0 | 0 |

### Supplementary table 7. Intra-familial comparisons of presence of risk alleles for the prioritized candidate SNVs

| | | Family 1 | | | Family 2 | | | Family 3 | | Family 4 | | Family 5 | | Family 6 | | Family 7 | | Family 8 | | Family 9 | | Family 10 | | Family 11 | |
|---|---|---|---|---|---|---|---|---|---|---|---|---|---|---|---|---|---|---|---|---|---|---|---|---|---|
| Gene ID | Variant ID | pG20D (FAST) | pG20D (noFAST) | Healthy | pG20D (noFAST) | Healthy | Healthy | *CLDN16* (noFAST) | Healthy | pG20D (noFAST) | Healthy | *CLDN19* (FAST) | *CLDN19* (FAST) | pG20D (FAST) | pG20D (noFAST) | pG20D (noFAST) | pG20D (noFAST) | *CLDN19* (noFAST) | *CLDN19* (noFAST) | *CLDN19* (noFAST) | *CLDN19* (noFAST) | pG20D (noFAST) | pG20D (noFAST) | *CLDN16* (noFAST) | *CLDN16* (noFAST) |
| CD86 | rs1129055 | | | | | | | | | | | | | | | | | | | | | | | | |
| CEP89 | rs3764633 | | | | | | | | | | | | | | | | | | | | | | | | |
| DMD | rs228406 | | | | | | | | | | | | | | | | | | | | | | | | |
| GALP | rs3745833 | | | | | | | | | | | | | | | | | | | | | | | | |
| IL12RB1 | rs401502 | | | | | | | | | | | | | | | | | | | | | | | | |
| IL12RB1 | rs375947 | | | | | | | | | | | | | | | | | | | | | | | | |
| IL12RB1 | rs11575934 | | | | | | | | | | | | | | | | | | | | | | | | |
| NFU1 | rs4453725 | | | | | | | | | | | | | | | | | | | | | | | | |
| SPATA20 | rs8065903 | | | | | | | | | | | | | | | | | | | | | | | | |
| AGBL2 | rs7941404 | | | | | | | | | | | | | | | | | | | | | | | | |
| ANKLE2 | rs7968520 | | | | | | | | | | | | | | | | | | | | | | | | |
| CLDN17 | rs35531957 | | | | | | | | | | | | | | | | | | | | | | | | |
| DMD | rs1800273 | | | | | | | | | | | | | | | | | | | | | | | | |
| HPS5 | rs7128017 | | | | | | | | | | | | | | | | | | | | | | | | |
| IL12RB1 | rs11575926 | | | | | | | | | | | | | | | | | | | | | | | | |
| ITGA1 | rs988574 | | | | | | | | | | | | | | | | | | | | | | | | |
| ZNF607 | rs61910733 | | | | | | | | | | | | | | | | | | | | | | | | |
| % Risk alleles | | 18% | 18% | 18% | 0% | 12% | 6% | 6% | 12% | 0% | 6% | 53% | 88% | 35% | 18% | 0% | 12% | 0% | 12% | 6% | 18% | 0% | 6% | 0% | 6% |
| % Non-risk alleles | | 82% | 82% | 82% | 100% | 88% | 94% | 94% | 88% | 100% | 94% | 47% | 12% | 65% | 82% | 100% | 88% | 100% | 88% | 94% | 82% | 100% | 94% | 100% | 94% |
| sum of risk alleles | | 3 | 3 | 3 | 0 | 2 | 1 | 1 | 2 | 0 | 1 | 9 | 15 | 6 | 3 | 0 | 2 | 0 | 2 | 1 | 3 | 0 | 2 | 0 | 2 |
